# Cost-Effectiveness of Platelet Function Testing in Dual Antiplatelet Therapy Decision-Making after Intracranial Aneurysm Treatment with Flow Diversion

**DOI:** 10.1101/2024.05.28.24308087

**Authors:** Aryan Wadhwa, Felipe Ramirez-Velandia, Emmanuel Mensah, Mira Salih, Alejandro Enriquez-Marulanda, Michael Young, Philipp Taussky, Christopher S. Ogilvy

## Abstract

**Introduction:** Dual antiplatelet therapy (DAPT) use is the standard of practice after flow diversion (FD) for intracranial aneurysms (IAs). Yet, no consensus exists in the literature regarding the optimal regimen. Certain institutions utilize various platelet function testing (PFT) to asses patient responsiveness to DAPT. Clopidogrel is the most commonly prescribed drug during DAPT, yet up to 52% of patients can be non-responders justifying PFT. Additionally, prices vary significantly among antiplatelet drugs, often further complicated by insurance restrictions. We aimed to determine the most cost-effective strategy for deciding DAPT regimens for patients after ICA treatment.

**Methods:** A decision tree with Monte Carlo simulations was performed to simulate patients undergoing various three-month postoperative DAPT regimens. Patients were either universally administered aspirin alongside Clopidogrel, Ticagrelor, or Prasugrel without PFT, or administered one of the former thienopyridine medications based on platelet reactivity unit (PRU) results after Clopidogrel. Input data for the model were extracted from the current literature, and the willingness-to-pay threshold (WTP) was defined as $100,000 per QALY as per standard practice in the US. The baseline comparison was with universal Clopidogrel DAPT without any PFT. Probabilistic and deterministic sensitivity analyses were performed to evaluate the robustness of the model.

**Results:** PFT-Prasugrel was the most cost-effective regimen compared to universal Clopidogrel, with a base-case incremental cost-effectiveness ratio (ICER) of $-35,255 (cost $2336.67, effectiveness 0.85). PFT-Ticagrelor (ICER $-4,671; cost $2,995.06, effectiveness 0.84), universal Prasugrel (ICER $5,553; cost $3,097.30, effectiveness 0.84), and universal Ticagrelor (ICER $75,969; cost $3,801.36, effectiveness 0.84) were all more cost-effective than treating patients with universal Clopidogrel (cost $3,041.77, effectiveness 0.83). These conclusions remain robust in probabilistic and deterministic sensitivity analyses.

**Conclusion:** The most cost-effective strategy for DAPT after FD for intracranial aneurysms is administering PFT-Prasugrel alongside aspirin. The cost of PFT is strongly justified and recommended when deciding patient-specific DAPT regimens.

## INTRODUCTION

Over the past decade, the adoption of Flow Diversion (FD) for the management of intracranial aneurysms (IAs) has shown consistent expansion.^1^ Flow diverters’ elevated metal coverage ratio necessitates the post-procedural application of dual antiplatelet therapy (DAPT) to mitigate the risk of thromboembolic complications (TECs) after stent placement. However, there is no consensus in the literature as to what the optimal postoperative DAPT regimen should be.

Across the US, the combination of Clopidogrel and aspirin represents the predominant choice for DAPT following FD.^2^ However, 16 to 52% of patients exhibit Clopidogrel resistance, potentially elevating their risk of TECs to as much as 17%.^3–5^ To address those concerns, up to 90% of US institutions now implement various platelet function testing (PFT) to determine patients’ responsiveness to Clopidogrel before FD stent placement.^2^ Currently, the VerifyNow test, which quantifies platelet reactivity units (PRU), stands as the prevailing PFT in neurointervention due to its practicality, allowing for bedside implementation.^3^

However, not only does PFT bear a cost, but alternative antiplatelet medications such as Prasugrel or Ticagrelor, which may be prescribed in cases of Clopidogrel resistance, are often more expensive for patients, potentially affecting compliance to DAPT.^6^ In some cases, health insurance companies are reluctant to approve higher costs for these alternative antiplatelet therapies unless PFT has been done.

This study aims to evaluate the cost-effectiveness of various strategies for determining the most appropriate postoperative DAPT regimen following FD intervention for unruptured IAs, focusing on comparing universally applied DAPT protocols and those guided by VerifyNow PFT outcomes.

## METHODS

### Setting

All data and information were obtained from literature sources. Due to the nature of these sources, approval from an institutional review board (IRB) was deemed unnecessary. This study did not receive any financial support for its conduct.

### Decision Tree

A decision tree model was built using TreeAge Pro 2024 (TreeAge Software, Inc.) to estimate the cost-effectiveness (measured by quality-adjusted life-years [QALYs]) of universal or PRU-driven use of DAPT in patients post-endovascular treatment for unruptured IAs. Post-procedural DAPT regimens reported in the included studies were analyzed, assuming all patients received appropriate preoperative loading doses of Clopidogrel. Our model assessed five different strategies, comprising: (1) universal and affordably priced Clopidogrel, control; (2) universal Ticagrelor; (3) universal Prasugrel; (4) PRU-guided Ticagrelor; and (5) PRU-guided Prasugrel. Double-dose Clopidogrel (150 mg/day) was omitted due to a lack of efficacy support from clinical trials.^7^ The universal strategies were created, utilizing reference data from cohorts with upfront use of thienopyridines without PFTs.^8,9^ In the Ticagrelor and Prasugrel PRU-guided treatment strategies, data was retrieved from meta-analyses and large cohorts, including patients undergoing VerifyNow P2Y12 prior to the procedure, guiding the need and change in therapy where appropriate.^10–12^ Patients included in these cohorts received Clopidogrel if documented to have adequate platelet inhibition or were switched to Ticagrelor or Prasugrel, depending on the clinician criteria. Maintenance dosages included in the studies were Clopidogrel 75 mg/day, Ticagrelor 90 mg twice daily, and Prasugrel 10 mg/day, each combined with aspirin (dose ranging from 75 to 325 mg/day, as appropriate).

The outcomes of the model were three different health states: (1) well after a thromboembolic or hemorrhagic complication (mRS of 0-2); (2) disabled after a thromboembolic or hemorrhagic complication (mRS 3-5); and (3) deceased after a thromboembolic or hemorrhagic complication. Utilizing a Monte Carlo simulation, parallel cohorts of 10,000 individual patients receiving DAPT after endovascular treatment for IAs were tracked across various regimens. The model started with the decision to initiate 1 of the 5 previously mentioned DAPT regimens. Each patient’s clinical pathway was simulated by sampling the probability distribution of relevant clinical parameters, including transition probabilities, utilities, and costs sourced from the literature. Costs, denominated in United States dollars (USD), and effectiveness, measured in QALYs, were then computed for comparison. The overall cost-effectiveness was assessed using net monetary benefit (NMB), which integrates cost, effectiveness, and the incremental cost-effectiveness ratio (ICER) to determine the optimal strategy. The ICER is defined as (cost of strategy 1 − cost of reference strategy)/(utility of strategy 1 − utility of reference strategy).

### Clinical Parameters

According to prior cost analyses, VerifyNow costs $30, and the yearly cost of a DAPT regimen with Clopidogrel is $639, Ticagrelor is $3,348, and Prasugrel is $2,496.^6^ Costs from disability, hemorrhage, and rates of thromboembolic complications (TECs) were retrieved from past cost analyses literature (**Table 1**).^13^

**Table 1.**
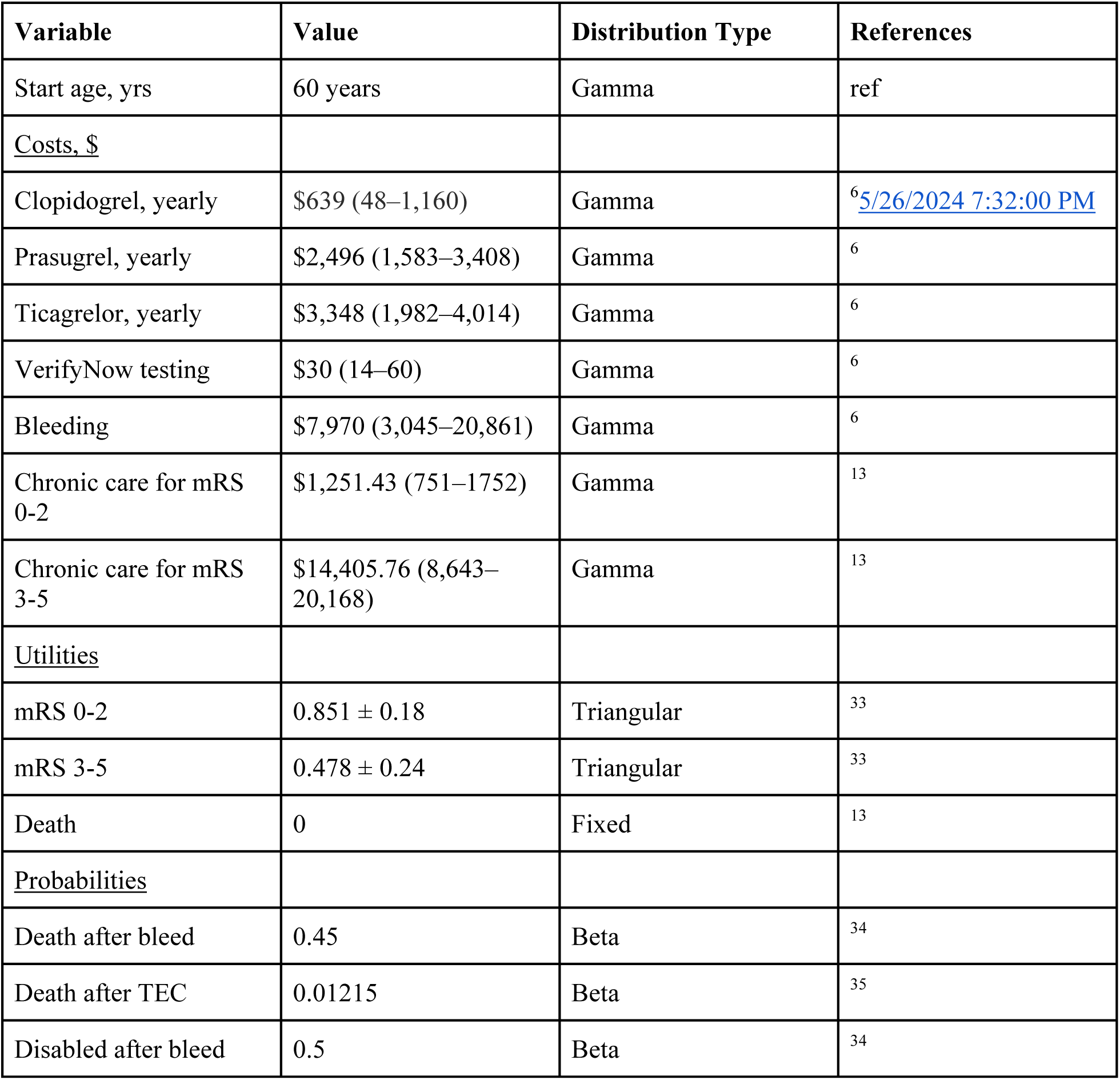
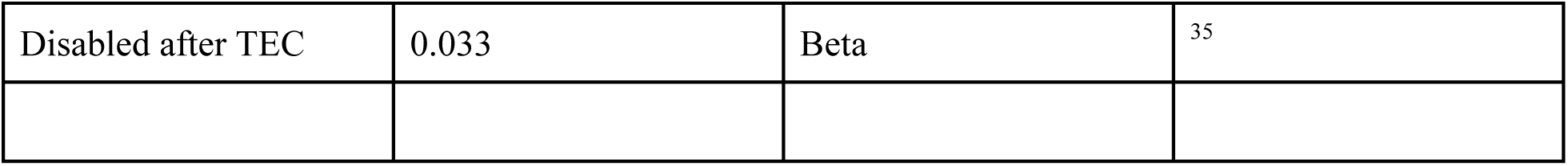
Clinical Parameters and Their Distribution.

| Variable | Value | Distribution Type | References |
| --- | --- | --- | --- |
| Start age, yrs | 60 years | Gamma | ref |
| <u>Costs, \$</u> | | | |
| Clopidogrel, yearly | \$639 (48–1,160) | Gamma | <sup>6</sup>5/26/2024 7:32:00 PM |
| Prasugrel, yearly | \$2,496 (1,583–3,408) | Gamma | <sup>6</sup> |
| Ticagrelor, yearly | \$3,348 (1,982–4,014) | Gamma | <sup>6</sup> |
| VerifyNow testing | \$30 (14–60) | Gamma | <sup>6</sup> |
| Bleeding | \$7,970 (3,045–20,861) | Gamma | <sup>6</sup> |
| Chronic care for mRS 0-2 | \$1,251.43 (751–1752) | Gamma | <sup>13</sup> |
| Chronic care for mRS 3-5 | \$14,405.76 (8,643–20,168) | Gamma | <sup>13</sup> |
| <u>Utilities</u> |  |  |  |
| mRS 0-2 | 0.851 ± 0.18 | Triangular | <sup>33</sup> |
| mRS 3-5 | 0.478 ± 0.24 | Triangular | <sup>33</sup> |
| Death | 0 | Fixed | <sup>13</sup> |
| <u>Probabilities</u> |  |  |  |
| Death after bleed | 0.45 | Beta | <sup>34</sup> |
| Death after TEC | 0.01215 | Beta | <sup>35</sup> |
| Disabled after bleed | 0.5 | Beta | <sup>34</sup> |
| Disabled after TEC | 0.033 | Beta | <sup>35</sup> |

All probability parameters were taken from the most up-to-date meta-analysis and large cohort studies (**Table 2**).^7–12^ Since the model was simulated over a short period of time (3 months), a discount rate and half-cycle correction was not deemed necessary to be applied to all the competing strategies. The willingness-to-pay (WTP) threshold was set at $100,000 per QALY gained as per the standard practice of cost-effectiveness analysis in the United States.

**Table 2.**
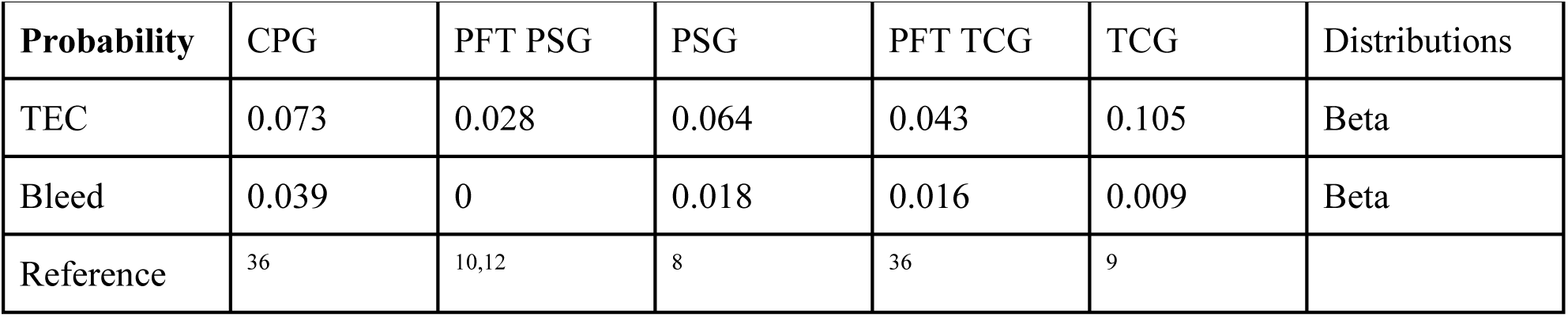
Rates of outcomes documented in the literature and used for the creation of the Decision Model.

| Probability | CPG | PFT PSG | PSG | PFT TCG | TCG | Distributions |
| --- | --- | --- | --- | --- | --- | --- |
| TEC | 0.073 | 0.028 | 0.064 | 0.043 | 0.105 | Beta |
| Bleed | 0.039 | 0 | 0.018 | 0.016 | 0.009 | Beta |
| Reference | <sup>36</sup> | <sup>10,12</sup> | <sup>8</sup> | <sup>36</sup> | <sup>9</sup> |  |

## Results

### Base Case Analysis

In the base case calculation, where mean values of the clinical parameters are utilized, PFT-Prasugrel yielded the highest cost-effectiveness of 0.85 QALYs with a total cost of $2,336.67 (ICER $-35,255; NMB $82,700). All other strategies were considered cost-effective compared to universal Clopidogrel, given that their ICERs were lower than the WTP, and all had higher net monetary benefits than universal Clopidogrel. The total costs and quality-adjusted life-years for each strategy during the 3-month simulated period are listed in **Table 3**.

**Table 3.** Base-case estimates of treatment costs and QALYs.

| Regimen | Costs | QALYs | ICER vs Universal Clopidogrel | NMB |
| --- | --- | --- | --- | --- |
| Universal Clopidogrel | \$3,041.77 | 0.83 | NA | \$79,834.57 |
| PFT-Prasugrel | \$2,336.67 | 0.85 | -35,255 | \$82,700.07 |
| Universal Prasugrel | \$3,097.30 | 0.84 | 5,553 | \$80,898.70 |
| PFT-Ticagrelor | \$2,995.06 | 0.84 | -4,671 | \$81,135.85 |
| Universal Ticagrelor | \$3,801.36 | 0.84 | 75,969 | \$80,602.73 |

### Deterministic Sensitivity Analysis

The deterministic sensitivity analysis involves adjusting the values of specific variables that may be relevant to the outcome, conducting this adjustment individually for one-way sensitivity analysis and in pairs for two-way sensitivity analysis, while maintaining the constancy of other variables. This process evaluates if the preferred strategy shifts when these variable ranges are altered. The variables noted to have the most considerable impact on incremental NMB were TEC and bleed mortality rates, along with disability rates after a TEC or bleed (**Figure 2A**). Given that variation in complication rates is the primary differentiating factor between regimens, other than cost, this comes as no surprise. An additional variable that we considered for a sensitivity analysis was the cost of platelet function testing itself, given that this would be a determining factor at many institutions (**Figure 2B**). Key parameters, such as treatment costs, effectiveness, and discount rates, were individually altered within plausible limits to evaluate their impact on the ICER. Threshold analyses identified the critical points at which parameter changes would alter the preferred strategy.

**Figure 1.**
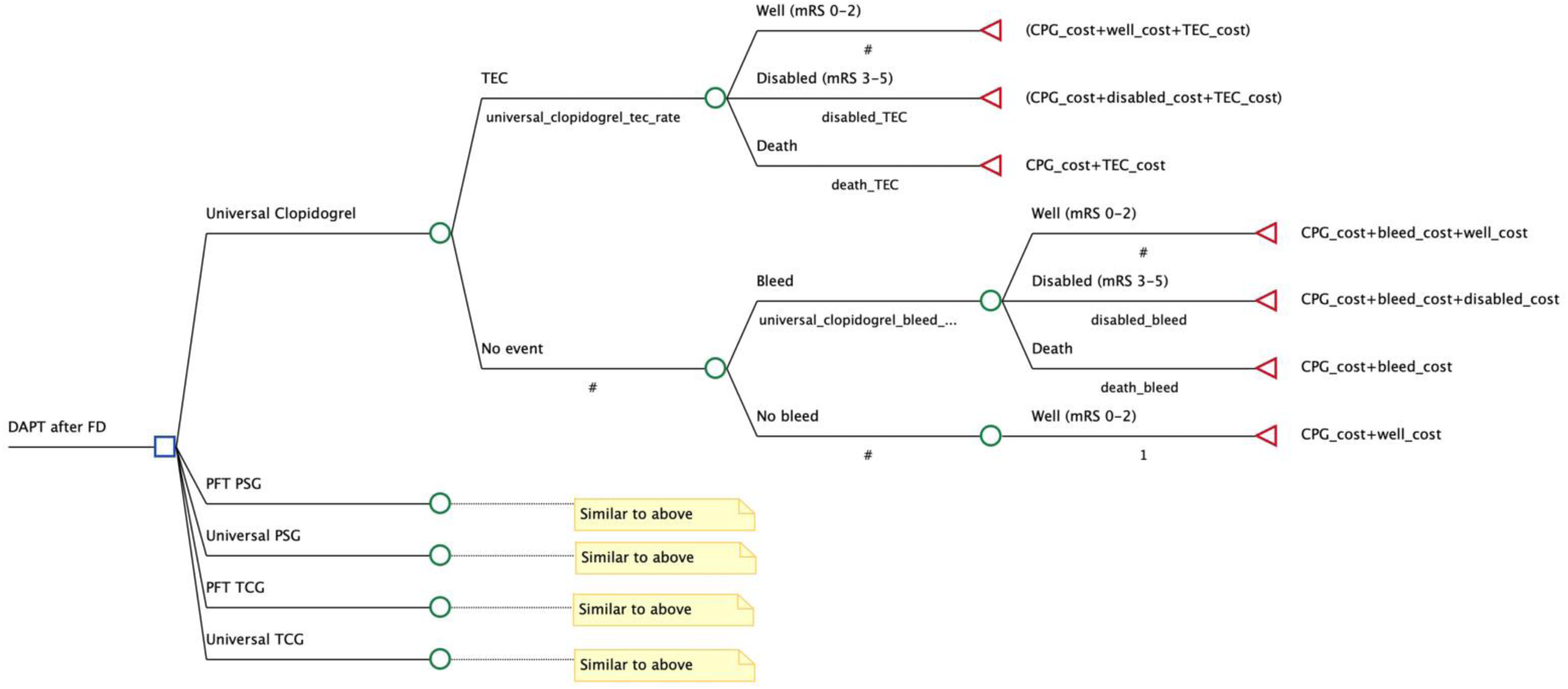
– Simplified decision tree used to model five different DAPT regimens after FD. TEC, thromboembolic complications. mRS, modified Rankin Scale.

**Figure 2A.**
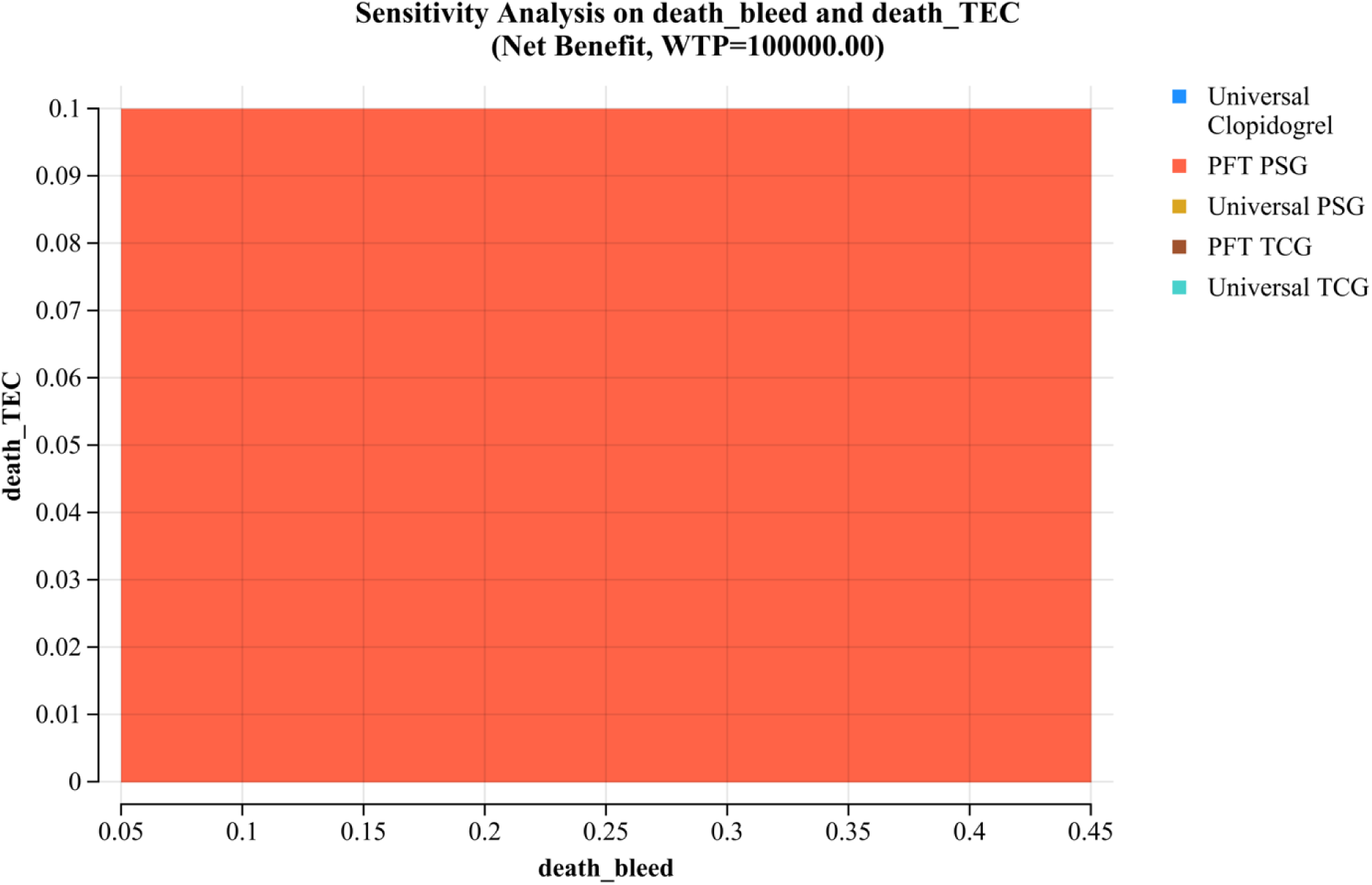
– Two-way sensitivity analysis varying the rate of mortality after a hemorrhagic complication (x-axis) and the rate of mortality after an ischemic complication (y-axis).

**Figure 2B.**
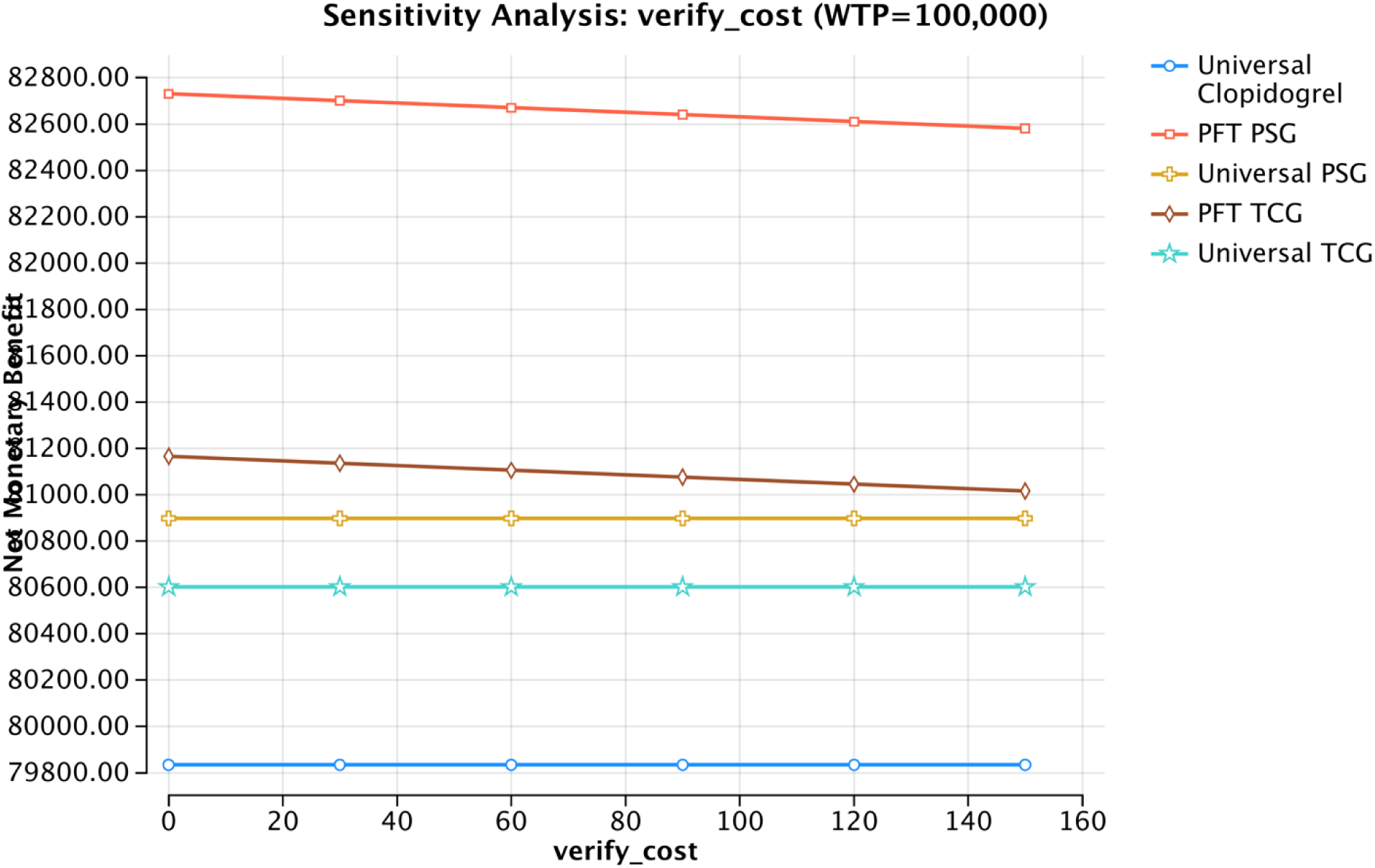
– One-way sensitivity analysis varying the cost of platelet function testing. A higher NMB is more cost-effective. The values on the x-axis and y-axis are USD.

### Probabilistic Sensitivity Analyses

A probabilistic sensitivity analysis was conducted, simulating a cohort of 10,000 patients across various iterations. Within this framework, five distinct follow-up strategies were evaluated using the Monte Carlo simulation method. This analysis revealed that PFT-PSG continued to be the most economically viable option, demonstrating the greatest NMB for any WTP threshold, as depicted in the cost-effectiveness acceptability curve (**Figure 3**). Further, Monte Carlo acceptability analysis indicated that PFT-PSG was identified as the most cost-effective strategy in approximately 70% of iterations, considering the standard WTP threshold of $100,000 per QALY (**Figure 3**).

**Figure 3.**
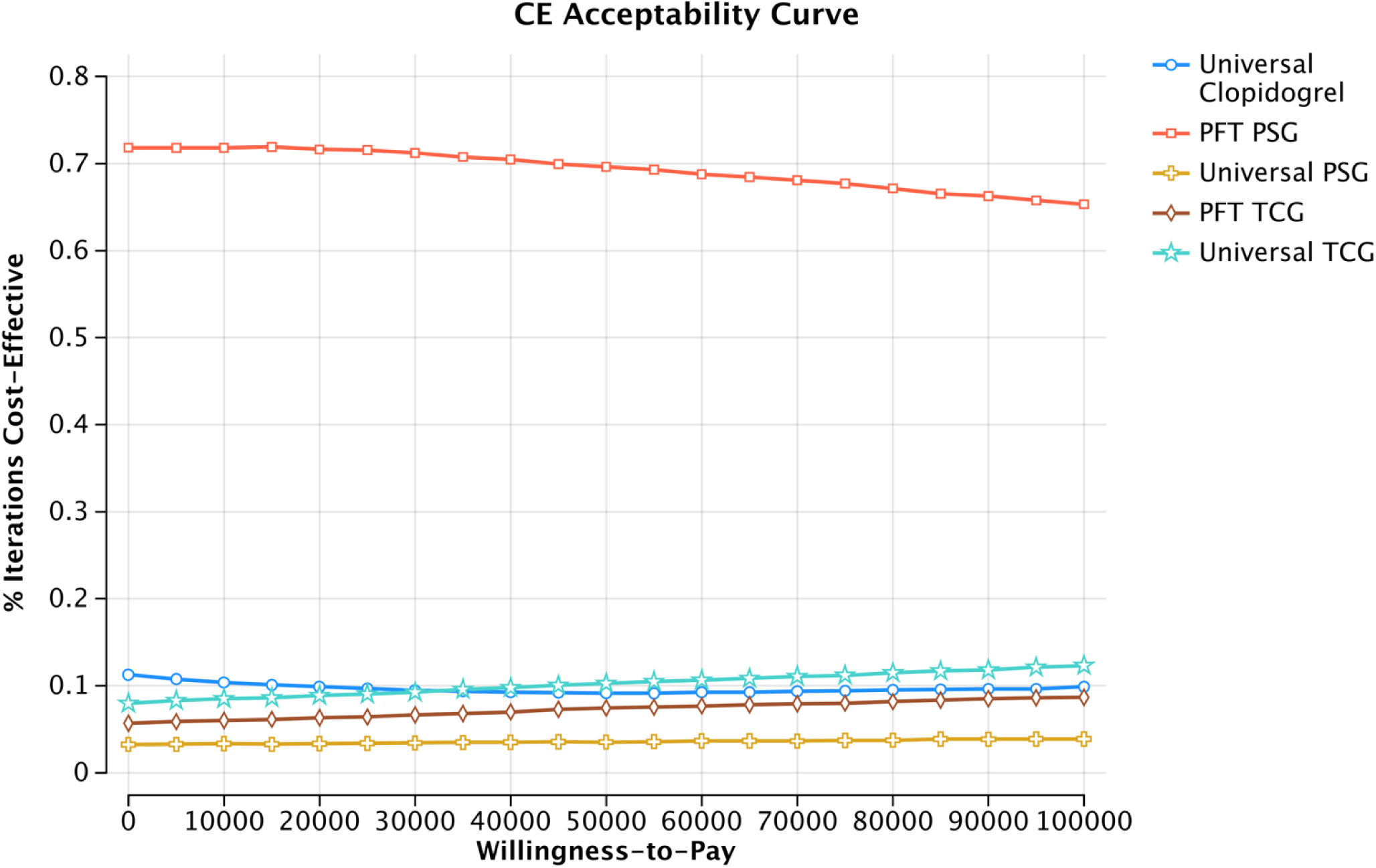
– Probabilistic sensitivity analysis: cost-effectiveness acceptability curve at various WTP values.

## Discussion

Cost-effective analyses, like our study utilizing a decision tree model, help understand the value of medical interventions in terms of cost per quality-adjusted life year (QALY) gained. Our study demonstrated that PFT-Prasugrel (Clopidogrel for responders and Prasugrel for non-responders detected on PFT) is the most cost-effective DAPT strategy for post-FD treatment of UIAs. Universal Prasugrel, PFT-Ticagrelor, and universal Ticagrelor were also considered cost-effective compared to universal Clopidogrel, assuming a WTP threshold of $100,000/QALY. Prasugrel and Ticagrelor are associated with higher annual drug costs, compared to Clopidogrel. However, these higher costs were offset in corresponding regimens by their better effectiveness in improving thromboembolic complications, which is critical in patients treated with FD. These findings conclude that the use of PFT to guide the choice of DAPT regimen following FD is not only clinically effective but also cost-effective.

Our study is the first of its kind in evaluating the cost-effectiveness of PFT to guide DAPT regimen choice for patients post-FD treatment for unruptured IAs. Previous studies in cardiovascular literature, however, have studied the cost-effectiveness of DAPT. Coleman et al. compared similar DAPT strategies in patients with acute coronary syndrome (ACS), demonstrating that platelet reactivity assay (PRA) (e.g., VerifyNow) driven Prasugrel and Ticagrelor were cost-effective to universal Clopidogrel.^6^ In this study, universal Ticagrelor and Prasugrel were also found to be cost-effective compared to their respective PRA-driven regimens. These results were most sensitive to differences in drug costs and drug-specific relative risks of death. It should be noted that whilst our study did not focus on a specific age group, Coleman et al. focused on a defined cohort of 65-year-old ACS patients.^6^ Additionally, the group’s economic analysis was primarily based on model assumptions and hypothetical cohorts, which might limit the generalizability of the results. Other cardiovascular studies have considered cost-effectiveness via genotype testing-driven DAPT regimens, checking for loss-of-function (LOF) alleles, after ACS and percutaneous coronary intervention. Limdi et al. found that utilizing both universal Ticagrelor and genotype-guided escalation of Ticagrelor was more cost-effective than universal Clopidogrel.^15^ A secondary analysis in this study, which evaluated genotype-guided de-escalation, showed higher effectiveness compared to nonguided de-escalation but was not cost-effective under most WTP thresholds. In addition to drug prices, the cost-effectiveness of genotype-guided strategies in this study could be sensitive to the prevalence of LOF genes. A second genotype-guided economic evaluation model by Kazi et al. showed genotype-guided therapy using Ticagrelor or Prasugrel for patients with LOF alleles is cost-effective compared to universal application of these drugs.^16^ In this study, genotype-guided Ticagrelor had an ICER of $30,200 per QALY compared to Clopidogrel. Temporal differences exist between our study and the evaluation by Kazi et al., who discuss lifetime costs and outcomes, compared to the shorter 3-month focus we used which is the typical amount of time that DAPT regimens are required post-FD.^16^ These studies evaluating the cost-effectiveness of DAPT regimens emphasize that personalized approaches to managing DAPT, whether through genetic testing or PFT, can lead to better economic and clinical outcomes. However, the specific focus on different patient populations, interventions, and biomarkers for personalization suggests that findings from each study are best applied within their respective contexts.

Despite our findings’ clear benefit of PFT, there is no consensus on the cut-off point for defining Clopidogrel hyporesponsiveness. Cut-off values of P2Y12 reaction units (PRU) of 85–208 have been described in cardiac literature as adequate Clopidogrel response, with PRU < 85 reported to be associated with a higher risk of bleeding events and PRU > 230 associated with significant morbidity and mortality.^6,17^ PRU studies in neurosurgery literature report adequate Clopidogrel response to fall between the ranges of PRU 60–240, with dose changes required at either end of the scale: for PRU < 60, lower doses are advised; for PRU >240, increasing Clopidogrel dose or switching to either Prasugrel or Ticagrelor is advised. To further reduce complication risk, additional Clopidogrel stratification has been employed, with Clopidogrel response defined as extreme response (PRU ≤ 15) and hyperresponse (PRU 16 – 60).^18^ In addition to Clopidogrel responsiveness, PRU responsiveness to Prasugrel has also been studied, with dose changes effected according to cut-off values. Higashiguchi et al. and Suyama et al. reported three different Prasugrel doses in patients post-FD treatment, with a 20 mg loading dose and daily 3.75mg for PRU > 210, daily 3.75mg for PRU 60 – 210, and daily 1.875 mg for PRU < 60.^8,19^ Variability in pharmacodynamic response to antiplatelets is also seen with Ticagrelor, where the established twice-daily 90 mg dose can lead to PRU values < 40, converting Clopidogrel hyporesponders to extreme responders.^18,20^

Additionally, universal Prasugrel was a cost-effective strategy and commonly preferred regimen at several institutions.^2^ Resistance to Prasugrel, which has been reported in other cohorts, has been more linked to medication adherence and pharmacological interactions, rather than genetic variants.^21^ In contrast to Clopidogrel, Prasugrel is a prodrug that requires activation through intestinal esterase and, to a lesser extent, CYP2C19 and CYP2C9.^22^ Additionally, unlike Clopidogrel, studies evaluating genetic polymorphisms have not revealed any effects on the pharmacokinetics and pharmacodynamic effects of Prasugrel.^23^ Possible explanations for this phenomenon include alternative activation through different hepatic CYP enzymes and higher potency as an irreversible ADP P2Y12 receptor antagonist.^3,24,25^ Due to Prasugrel’s high potency, some initial concerns arose regarding the heightened risk of intracranial hemorrhage in patients with a history of ischemic stroke.^26,27^ For outpatient neurovascular procedures, as unruptured IAs treated with FD, various cohorts have demonstrated a favorable safety profile for Prasugrel.^28–31^ In the meta-analysis by Podlasek et al., comprising 49 studies and 2,526 patients undergoing FD treatment for unruptured IAs, hemorrhagic events were negligible for Prasugrel.^12^ Similarly, in the cohort reported by Souyama et al., in which there was routine use of Prasugrel post-FD for 3-6 months, out of the 110 patients, only one had a hemorrhage after treatment.^8^ Overall, these findings underscore Prasugrel’s promising safety and efficacy profile in the context of FD, warranting further exploration in prospective clinical trials.

While insightful, our study is not without limitations. The reliance on retrospective cohorts and model-based simulations may not capture the full complexity of individual patient responses in a real-world setting. The heterogeneity of included studies also limits our retrospective approach in determining thromboembolic and hemorrhagic rates. The paucity of data from meta-analyses also led to our reliance on the best available retrospective cohorts, which has limitations. Retrospective analyses are subject to selection bias and confounding. This, along with the heterogeneity in the sample set and variation in the duration of DAPT, may produce findings that are not always generalizable. Moreover, we also omitted double-dose Clopidogrel as a DAPT regimen, which may be efficacious in a subset of individuals with Clopidogrel hyporesponsiveness.^32^ An additional limitation that was difficult to circumvent due to the lack of stratified reporting in the literature was the difference in the rates of various types of bleeding events and the mortality rates of each of those types, which also varies by type of regimen. There were not enough studies considering all these possible variations and elucidating each of these individual rates. Finally, our use of the decision tree model for economic valuations, which is contingent on the accuracy of the cost data and assumptions we embedded in the model, might not reflect the dynamic nature of healthcare economics and patient management. Notably, our decision tree model did not account for the influence of age, gender, or specific patient comorbidities. It also assumed that rates of thromboembolic and hemorrhagic events would remain constant over the three-month study period. Thus, future studies should aim to incorporate more diverse patient data and real-world outcomes, where available, to validate and refine the proposed cost-effectiveness framework.

## Conclusion

We have demonstrated that PFT-guided therapy, using Prasugrel for Clopidogrel hyporesponsiveness, was the most cost-effective strategy guiding DAPT post-FD treatment of unruptured IAs. Furthermore, prescribing universal Prasugrel, PFT-guided Ticagrelor, or universal Ticagrelor were also cost-effective compared to prescribing universal Clopidogrel. Although this sheds light on the potential cost benefits of initiating Prasugrel DAPT upfront, future prospective, and ideally randomized clinical trials are necessary to evaluate the safety of this intervention post-FD.

## Declarations

### Availability of data and material

All information in the study is publicly available

### Code Availability

No code were used in this study

### Ethics approval

Due to the nature of this study being a retrospective analysis of publicly available data, IRB approval was not deemed necessary for this study.

### Consent to participate

Informed consent not required due to the nature of this study.

### Consent for publication

Authors provide consent for publicationof all materials

## Data Availability

All data in this manuscript was found from literature publicly available, and referenced in the manuscript.

## Acknowledgements

The study team would like to thank the Brain Aneurysm Institute. Their invaluable support has been crucial in providing comprehensive care to aneurysm patients and contributing to the research of intracranial aneurysms.

## References

1. Mirpuri P, Khalid SI, McGuire LS, Alaraj A. Trends in Ruptured and Unruptured Aneurysmal Treatment from 2010 to 2020: A Focus on Flow Diversion. World Neurosurg. 2023;178:e48–e56. doi:10.1016/j.wneu.2023.06.093

2. Gupta R, Moore JM, Griessenauer CJ, et al. Assessment of Dual-Antiplatelet Regimen for Pipeline Embolization Device Placement: A Survey of Major Academic Neurovascular Centers in the United States. World Neurosurg. 2016;96:285–292. doi:10.1016/j.wneu.2016.09.013

3. Taylor LI, Dickerson JC, Dambrino RJ, et al. Platelet testing in flow diversion: a review of the evidence. Neurosurg Focus. 2017;42(6):E5. doi:10.3171/2017.3.FOCUS1746

4. Lim ST, Thijs V, Murphy SJX, et al. Platelet function/reactivity testing and prediction of risk of recurrent vascular events and outcomes after TIA or ischaemic stroke: systematic review and meta-analysis. J Neurol. 2020;267(10):3021–3037. doi:10.1007/s00415-020-09932-y

5. Adeeb N, Griessenauer CJ, Foreman PM, et al. Use of Platelet Function Testing Before Pipeline Embolization Device Placement: A Multicenter Cohort Study. Stroke. 2017;48(5):1322–1330. doi:10.1161/STROKEAHA.116.015308

6. Coleman CI, Limone BL. Cost-Effectiveness of Universal and Platelet Reactivity Assay-Driven Antiplatelet Therapy in Acute Coronary Syndrome. The American Journal of Cardiology. 2013;112(3):355–362. doi:10.1016/j.amjcard.2013.03.036

7. Mehta SR, Tanguay JF, Eikelboom JW, et al. Double-dose versus standard-dose clopidogrel and high-dose versus low-dose aspirin in individuals undergoing percutaneous coronary intervention for acute coronary syndromes (CURRENT-OASIS 7): a randomised factorial trial. Lancet. 2010;376(9748):1233–1243. doi:10.1016/S0140-6736(10)61088-4

8. Suyama K, Nakahara I, Matsumoto S, et al. Efficacy and Safety of Dual Antiplatelet Therapy with the Routine Use of Prasugrel for Flow Diversion of Cerebral Unruptured Aneurysms. Clin Neuroradiol. 2024;34(1):201–208. doi:10.1007/s00062-023-01355-2

9. Papaxanthos J, Cagnazzo F, Collemiche FL, et al. Ticagrelor versus clopidogrel dual antiplatelet therapy for unruptured intracranial aneurysms treated with flowdiverter. J Neuroradiol. 2023;50(3):346–351. doi:10.1016/j.neurad.2022.11.010

10. Flynn LM, Mohamed E, Dobbs N, et al. Safety of dual antiplatelet therapy using aspirin and low-dose Prasugrel with platelet reactivity testing in flow diverter treatment of intracranial aneurysms. Interv Neuroradiol. Published online November 29, 2023:15910199231217142. doi:10.1177/15910199231217142

11. Neyens R, Donaldson C, Andrews C, Kellogg R, Spiotta A. Platelet Function Testing with a VerifyNow-Directed Personalized Antiplatelet Strategy and Associated Rates of Thromboembolic Complications After Pipeline Embolization for Complex Cerebral Aneurysms. World Neurosurg. 2020;138:e674–e682. doi:10.1016/j.wneu.2020.03.046

12. Podlasek A, Al Sultan AA, Assis Z, Kashani N, Goyal M, Almekhlafi MA. Outcome of intracranial flow diversion according to the antiplatelet regimen used: a systematic review and meta-analysis. J Neurointerv Surg. 2020;12(2):148–155. doi:10.1136/neurintsurg-2019-014996

13. Salem MM, Salih M, Nwajei F, et al. Cost-Effectiveness Analytic Comparison of Neuroimaging Follow-Up Strategies After Pipeline Embolization Device Treatment of Unruptured Intracranial Aneurysms. World Neurosurg. 2022;158:e206–e213. doi:10.1016/j.wneu.2021.10.154

14. Arias E, Xu J, Kochanek K. United States Life Tables, 2021. Natl Vital Stat Rep. 2023;72(12):1–64.

15. Limdi NA, Cavallari LH, Lee CR, et al. Cost-effectiveness of CYP2C19-guided antiplatelet therapy in patients with acute coronary syndrome and percutaneous coronary intervention informed by real-world data. Pharmacogenomics J. 2020;20(5):724–735. doi:10.1038/s41397-020-0162-5

16. Kazi DS, Garber AM, Shah RU, et al. Cost-effectiveness of genotype-guided and dual antiplatelet therapies in acute coronary syndrome. Ann Intern Med. 2014;160(4):221–232. doi:10.7326/M13-1999

17. Sibbing D, Aradi D, Alexopoulos D, et al. Updated Expert Consensus Statement on Platelet Function and Genetic Testing for Guiding P2Y12 Receptor Inhibitor Treatment in Percutaneous Coronary Intervention. JACC Cardiovasc Interv. 2019;12(16):1521–1537. doi:10.1016/j.jcin.2019.03.034

18. Young CC, Bass DI, Cruz MJ, et al. Clopidogrel hyper-response increases peripheral hemorrhagic complications without increasing intracranial complications in endovascular aneurysm treatments requiring dual antiplatelet therapy. J Clin Neurosci. 2022;105:66–72. doi:10.1016/j.jocn.2022.09.005

19. Higashiguchi S, Sadato A, Nakahara I, et al. Reduction of thromboembolic complications during the endovascular treatment of unruptured aneurysms by employing a tailored dual antiplatelet regimen using aspirin and prasugrel. J Neurointerv Surg. 2021;13(11):1044–1048. doi:10.1136/neurintsurg-2020-016994

20. Kim KS, Fraser JF, Grupke S, Cook AM. Management of antiplatelet therapy in patients undergoing neuroendovascular procedures. J Neurosurg. 2018;129(4):890–905. doi:10.3171/2017.5.JNS162307

21. Lhermusier T, Lipinski MJ, Tantry US, et al. Meta-analysis of direct and indirect comparison of ticagrelor and prasugrel effects on platelet reactivity. Am J Cardiol. 2015;115(6):716–723. doi:10.1016/j.amjcard.2014.12.029

22. Krishnan K, Nguyen TN, Appleton JP, et al. Antiplatelet Resistance: A Review of Concepts, Mechanisms, and Implications for Management in Acute Ischemic Stroke and Transient Ischemic Attack. Stroke: Vascular and Interventional Neurology. 2023;3(3):e000576. doi:10.1161/SVIN.122.000576

23. Brandt JT, Close SL, Iturria SJ, et al. Common polymorphisms of CYP2C19 and CYP2C9 affect the pharmacokinetic and pharmacodynamic response to clopidogrel but not prasugrel. J Thromb Haemost. 2007;5(12):2429–2436. doi:10.1111/j.1538-7836.2007.02775.x

24. Mega JL, Close SL, Wiviott SD, et al. Cytochrome P450 genetic polymorphisms and the response to prasugrel: relationship to pharmacokinetic, pharmacodynamic, and clinical outcomes. Circulation. 2009;119(19):2553–2560. doi:10.1161/CIRCULATIONAHA.109.851949

25. Flechtenmacher N, Kämmerer F, Dittmer R, et al. Clopidogrel Resistance in Neurovascular Stenting: Correlations between Light Transmission Aggregometry, VerifyNow, and the Multiplate. AJNR Am J Neuroradiol. 2015;36(10):1953–1958. doi:10.3174/ajnr.A4388

26. Akbari SH, Reynolds MR, Kadkhodayan Y, Cross DT, Moran CJ. Hemorrhagic complications after prasugrel (Effient) therapy for vascular neurointerventional procedures. J Neurointerv Surg. 2013;5(4):337–343. doi:10.1136/neurintsurg-2012-010334

27. Kitazono T, Toyoda K, Kitagawa K, et al. Efficacy and Safety of Prasugrel by Stroke Subtype: A Sub-Analysis of the PRASTRO-I Randomized Controlled Trial. J Atheroscler Thromb. 2021;28(2):169–180. doi:10.5551/jat.56093

28. Chalouhi N, Tjoumakaris S, Phillips JLH, et al. A single pipeline embolization device is sufficient for treatment of intracranial aneurysms. AJNR Am J Neuroradiol. 2014;35(8):1562–1566. doi:10.3174/ajnr.A3957

29. Stetler WR, Chaudhary N, Thompson BG, Gemmete JJ, Maher CO, Pandey AS. Prasugrel is effective and safe for neurointerventional procedures. J Neurointerv Surg. 2013;5(4):332–336. doi:10.1136/neurintsurg-2012-010302

30. Daou B, Starke RM, Chalouhi N, et al. P2Y12 Reaction Units: Effect on Hemorrhagic and Thromboembolic Complications in Patients With Cerebral Aneurysms Treated With the Pipeline Embolization Device. Neurosurgery. 2016;78(1):27. doi:10.1227/NEU.0000000000000978

31. Guédon A, Clarençon F, Maria FD, et al. Very late ischemic complications in flow-diverter stents: a retrospective analysis of a single-center series. Journal of Neurosurgery. 2016;125(4):929–935. doi:10.3171/2015.10.JNS15703

32. Sardella G, Calcagno S, Mancone M, et al. Pharmacodynamic effect of switching therapy in patients with high on-treatment platelet reactivity and genotype variation with high clopidogrel Dose versus prasugrel: the RESET GENE trial. Circ Cardiovasc Interv. 2012;5(5):698–704. doi:10.1161/CIRCINTERVENTIONS.112.972463

33. Wali AR, Park CC, Santiago-Dieppa DR, Vaida F, Murphy JD, Khalessi AA. Pipeline embolization device versus coiling for the treatment of large and giant unruptured intracranial aneurysms: a cost-effectiveness analysis. Neurosurg Focus. 2017;42(6):E6. doi:10.3171/2017.3.FOCUS1749

34. Brinjikji W, Lanzino G, Cloft HJ, Siddiqui AH, Kallmes DF. Risk Factors for Hemorrhagic Complications following Pipeline Embolization Device Treatment of Intracranial Aneurysms: Results from the International Retrospective Study of the Pipeline Embolization Device. AJNR Am J Neuroradiol. 2015;36(12):2308–2313. doi:10.3174/ajnr.A4443

35. Brinjikji W, Lanzino G, Cloft HJ, et al. Risk Factors for Ischemic Complications following Pipeline Embolization Device Treatment of Intracranial Aneurysms: Results from the IntrePED Study. AJNR Am J Neuroradiol. 2016;37(9):1673–1678. doi:10.3174/ajnr.A4807

36. Zhou Y, He S, Hu Y. One year clinical outcome of dual anti-platelet therapy with the Novel Ticagrelor plus Aspirin versus Clopidogrel plus Aspirin for Endovascular Intervention of patients with Intracranial Aneurysm: A meta-analysis. Journal of Stroke and Cerebrovascular Diseases. 2024;33(1):107491. doi:10.1016/j.jstrokecerebrovasdis.2023.107491

